# Integration of Elemental Imaging and Spatial Transcriptomic Profiling for Proof-of-Concept Metals-Based Pathway Analysis of Colon Tumor Microenvironment

**DOI:** 10.1101/2024.12.09.24318747

**Authors:** Aruesha Srivastava, Neha Shaik, Yunrui Lu, Matthew Chan, Alos Diallo, Serin Han, Tracy Punshon, Brian Jackson, Linda Vahdat, Xiaoying Liu, Vivek Mittal, Ken Lau, Jiang Gui, Louis Vaickus, Jack Hoopes, Fred Kolling, Laurent Perreard, Jonathan Marotti, Joshua Levy

## Abstract

The complex interplay between metal abundance, transport mechanisms, cell distribution, and tumor progression-related biological pathways (e.g., metabolism, collagen remodeling) remains poorly understood. Traditionally, genes and metals have been studied in isolation, limiting insights into their interactions. Recent advances in spatial transcriptomics and elemental profiling now enable comprehensive exploration of tissue-wide metal-gene interactions, though integration remains challenging. In this proof-of-concept study, we investigated metal-dependent signaling within the tumor microenvironment of a unique colorectal cancer (CRC) tumor. We implemented a spatial multimodal workflow which integrated elemental imaging, gene expression, cellular composition, and histopathological features to uncover metals-related pathways through spatially resolved differential expression analysis. Preliminary findings revealed significant associations, for instance: elevated iron correlated with mesenchymal phenotypes located at the tumor’s proliferative front, reflecting epithelial-to-mesenchymal transition pathways, and extracellular matrix remodeling. High concentrations of copper were predominantly localized in regions of active tumor growth and associated with the upregulation of immune response genes. This proof-of-concept workflow demonstrates the feasibility of integrating elemental imaging with spatial transcriptomics to identify metals-based gene correlates. Future application of this workflow to larger patient cohorts will pave the way for expansive comparisons across the metallome and transcriptome, ultimately identifying novel targets for tumor progression biomarkers and therapeutic interventions.

## Background and Introduction

Colorectal Cancer (CRC) represents a significant global health challenge, accounting for nearly 10% of all cancer cases and ranking as the second-leading cause of cancer-related deaths worldwide ^1^. The rising incidence of CRC among younger demographics underscores an urgent need to advance screening, prognostic tools, and therapeutic approaches ^2,3^. Central to improving outcomes is a deeper understanding of the mechanisms underlying tumor progression and metastasis, which are responsible for approximately 90% of cancer mortalities and associated with sharply declining survival rates at advanced stages.

Elements significantly influence cancer progression through roles in cell proliferation, invasion, motility, adhesion, and more ^4–6^. Metals such as copper (Cu), iron (Fe), and zinc (Zn) are vital for enzymatic reactions essential for mitochondrial respiration, DNA repair, senescence, and immune regulation ^7–18^. Metals also modulate signaling pathways through metalloallostery, influencing nutrient sensing and protein regulation. Elevated levels of Cu and Fe can also contribute to reactive oxygen species (ROS) production, which promotes angiogenesis and disrupts DNA repair, enhancing tumor invasion and metastasis ^15,19,20^. The interplay between various essential and non-essential elements is integral to tumor growth and metastasis, and research into these interactions and their biological function will provide insights into element-dependent vulnerabilities, offering potential targets for novel therapeutic interventions. For instance, a recent Phase II trial with tetrathiomolybdate (TM), a Cu chelator, demonstrated promising results in improving progression-free and overall survival rates for breast cancer patients at high risk of metastasis ^21–23^. TPEN, another Cu chelator, selectively targets CRC cells due to their higher Cu accumulation ^12,17^. Despite advances in metal-based diagnostics and therapies, their clinical application faces significant challenges due to incomplete understanding of how metals are distributed within tumors and their specific roles in intra- and inter-cellular signaling within the tumor microenvironment (TME). The complexity of elemental distribution, influenced by factors such as dietary intake and unique cellular uptake and export mechanisms, results in a dynamic and heterogeneous metal landscape within tumors ^24–26^. This complexity is further exacerbated by the presence of metals in various cellular pools, ranging from tightly bound to more labile forms that interact with small molecules ^27^.

A deeper understanding of metal distribution within tumors has been traditionally limited by bulk analysis methods, which overlook the nuanced spatial distribution of metals crucial for understanding their role in cancer pathogenesis. High-resolution elemental imaging (EI) such as Laser Ablation Inductively Coupled Plasma Time of Flight Mass Spectrometry (LA-ICP-TOF-MS) offers a significant breakthrough by providing precise localization of metal accumulation.

This technology enables detailed mapping of metal pools within the tumor landscape, revealing its potential influence on cancer progression and patient outcomes. It holds the potential to revolutionize cancer diagnosis and treatment by identifying unique multi-element signatures indicative of tumor states.

Complementing this, recent technological advances in spatial transcriptomics (ST) enable high- resolution ^28^, untargeted spatial expression profiling of almost all protein-coding genes, providing insights into the genetic factors governing metal transport and their toxicological implications ^29^. The conserved nature of metal transport proteins across species underscores the relevance of these genetic studies. ST overcomes the limitations of bulk analysis, which can obscure tissue-specific relationships, and of multiplexing assays, which are restricted to a limited number of protein candidates ^30–38^. By allowing for the profiling of the entire transcriptome at high spatial resolution, including at the single-cell level, ST can potentially be leveraged to pinpoint specific cellular interactions or markers of tumor progression.

The spatial location and abundance of essential elements within tissues reflect complex processes of availability, homeostasis, and biological necessity. For instance, the homeostasis of essential elements involves a myriad of proteins that sense, signal, chaperone, and control their movement ^39,40^. Thus, integrating spatial transcriptomics and elemental imaging technologies has the potential to reveal intricate metal-biomolecular interactions that may be missed by traditional reductionist approaches that would analyze each assay in isolation ^41^. Yet, the absence of dedicated software platforms specifically designed for such integrative tasks has been a significant obstacle, explaining, in part, why comprehensive pathway analysis for metals in cancer remains an unrealized goal to date ^42–45^.

To address this gap, the Biomedical National Elemental Imaging Resource (BNEIR) developed TRACE (Tissue Region Analysis through Co-registration of Elemental Maps), co-registration software that facilitates the spatial integration of elemental imaging data with histopathology, immunohistochemistry/multiplex immunofluorescence, and spatial transcriptomics technologies (**Figure 1**) ^41,46,47^. This exploratory proof-of-concept study builds on TRACE-enabled integration to further characterize elemental and transcriptomic data from the primary tumor of a single colorectal cancer (CRC) case.

**Figure 1:**
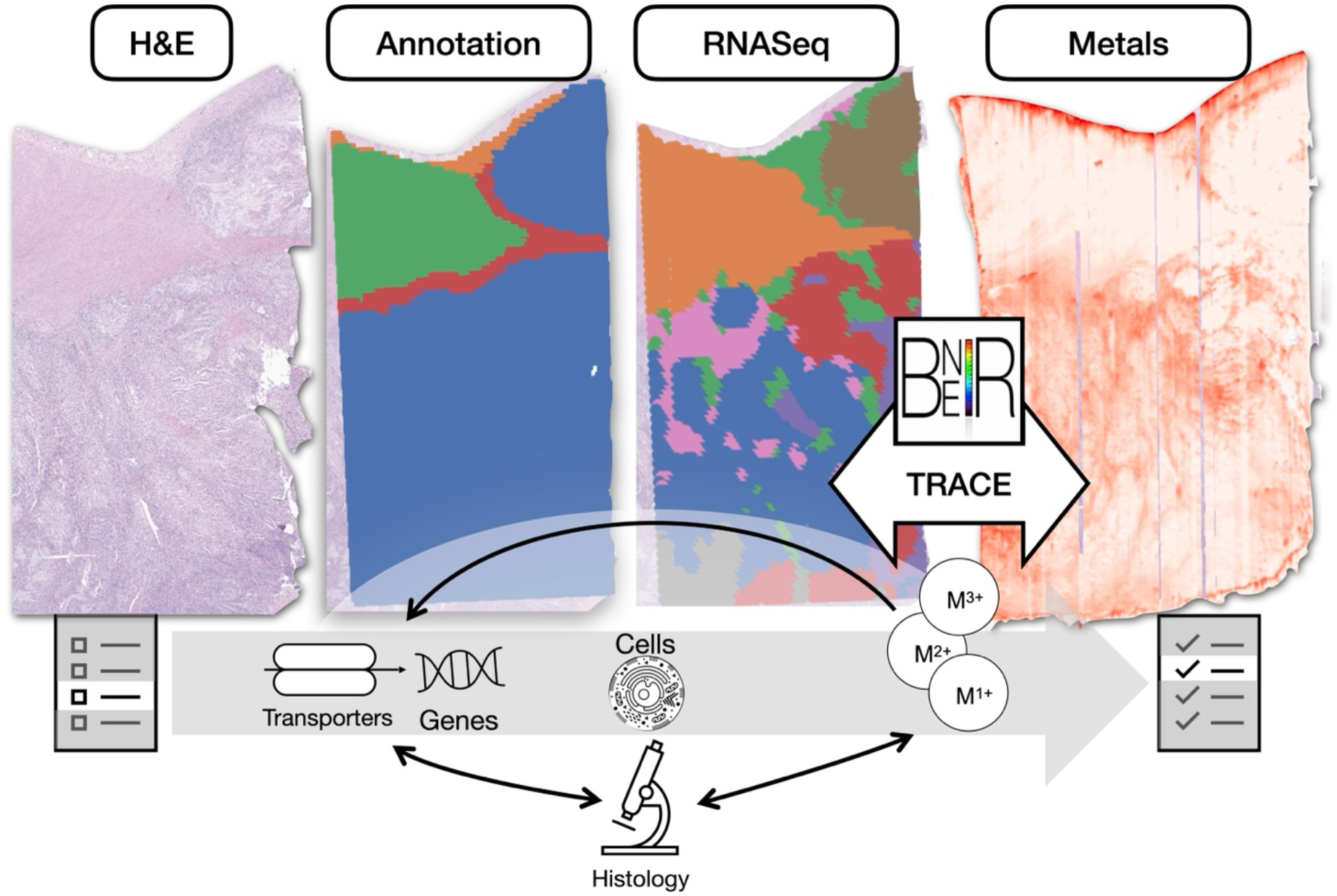
Overview: Spatial Integration of Spatial Elemental Imaging and Spatial Transcriptomics can reveal genes associated with metal bioaccumulation within specific tissue architectures, shedding light on metals-related pathways and cellular changes associated with tumorigenesis; BNEIR: Biomedical National Elemental Imaging Resource; TRACE: Tissue Region Analysis through Co-registration of Elemental Maps

Through a multimodal analysis, we aim to demonstrate the potential for uncovering valuable insights into the interplay between genetic and elemental landscapes in cancer pathology, paving the way for a more comprehensive understanding of CRC progression. The primary objectives of this study are twofold: firstly, to identify correlations between gene signatures and metal abundance within specific cellular architectures and cell types in CRC; secondly, to leverage these insights to develop a metals-based pathway analysis. We believe this initial investigation will facilitate a more comprehensive exploration of metal-related pathways in cancer across larger cohorts, enhancing our understanding of pathogenesis, metastasis, and progression (**Figure 1**). Ultimately, this research seeks to clarify the role of metal bioaccumulation in tumor dynamics, potentially leading to the discovery of novel biomarkers and the development of more effective therapeutic strategies.

## Results

### Results Overview

We conducted a comprehensive “metals-based pathway analysis” on a primary tumor section from a CRC patient, aiming to uncover associations between the abundance of various metals and gene pathways within distinct tissue architectures. The primary tumor, originating in the patient’s cecum at pathologic T-stage 3 (indicating invasion through the muscularis propria), advanced to stage 2a lymph node involvement and metastasized to the liver. This section provides a concise overview as context for the subsequent findings:

1. **Spatial Transcriptomics (ST) Profiling:** Utilized the 10x Genomics Visium spatial transcriptomic (ST) CytAssist assay to capture spatial variations in the expression of approximately 18,000 genes across 55-micron spots. This was complemented by high-resolution 40X H&E-stained whole slide imaging (WSI; Leica Aperio GT450) on the same section.
2. **Spatial Elemental Imaging (EI):** A serial section was analyzed to profile all elements and their isotopes at 5-micron resolution using laser ablation inductively coupled plasma time-of-flight mass spectrometry (LA-ICPTOF-MS).
3. **Spatial Data Integration:** Achieved through TRACE (Tissue Region Analysis through Co-registration of Elemental Maps), which facilitated the spatial alignment of ST and EI data.
4. **Pathologist annotations:** Annotated WSI identified regions inside, around, and away from the tumor, among other tissue architectures such as epithelium, serosa, and subcutaneous fat.
5. **Elemental Hotspot analysis:** Identifies hotspot areas of high and low metal abundance using Getis Ord* statistics ^48^.
6. **Cell Typing:** Integrated single cell RNASeq data from a serial section to characterize cell types within these hotspots.
7. **Differential Expression:** Conducted a transcriptome-wide comparison of gene expression in areas with varying metal abundance.
8. **Pathway Analysis:** Gene set enrichment analysis performed on statistically significant genes to elucidate the connections between metal abundance and various biological processes. Metal-gene correlations were also visualized across genes contained within several select, relevant Cu homeostasis and tumor progression pathways for additional context, as an example of how spatial data integration can recapitulate and expand on known biological mechanisms and pathways.
9. **Factor/Interaction Analysis:** Employed machine learning and clustering approaches to reveal distinct profiles of metals, genes, and cell types associated with different tissue histologies.

### Identifying tissue architectural and cellular components associated with high elemental abundance

Our hotspot analysis revealed distinct elemental signatures associated with various histological structures (**Figures 2A-C, S1, Tables S1-2**). Notably, the tumor regions were enriched in Cu, Mg, Fe, and Mn. These differences were accentuated with respect to other tissue regions such as the muscularis propria. Conversely, K, Zn, were relatively depleted in the tumor compared to the muscularis propria. Zn concentrations were particularly high at the tumor interface.

**Figure 2:**
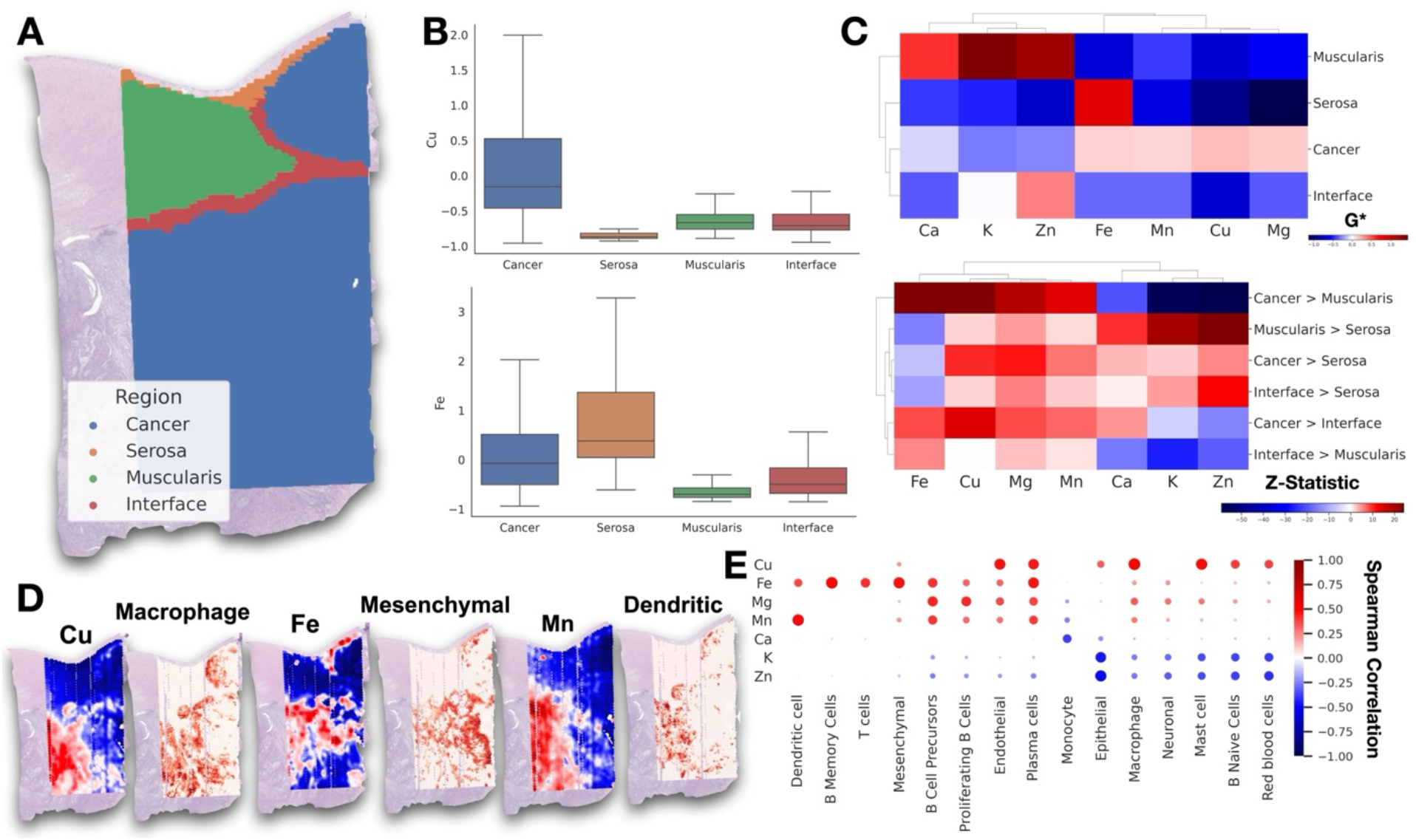
Metal distribution in tissue architectures: **A)** Pathologist annotation of tumor, tumor interface, and surrounding musculature and serosa, **B)** Boxplot demonstrating differences in Gi* hotspot statistics across various tissue architectures, **C)** Clustered heatmaps comparing average hotspot abundance (Gi*) by metal within select architectures and relative differences in hotspot abundance between architectures (positive z-score indicates higher metal abundance on architecture to left of inequality), **D)** Demonstration of metal abundance mapped across slide for Fe and Mn (red indicates hotspot, blue indicates coldspot), juxtaposed with cellular abundance (red indicates higher abundance), **E)** Dotplot demonstrating spearman correlation associations between cell type abundance and elemental distribution– larger red dots indicate positive associations of greater magnitude, whereas blue dots indicate negative associations, with size indicating magnitude; “Inter” represents the tumor interface

Additionally, the muscularis propria showed significant enrichments of metals such as Zn, K, Ca as compared to serosal tissue.

Deconvolution of ST into cell-type proportions revealed significant associations between various metals and specific cell types (**Figures 2D,E, S1, Table S3**). For instance, Cu was positively associated with presence of mast cells, B naive cells, endothelial cells, macrophages, and plasma cells, indicating a broad involvement across immune and vascular functions. Fe was associated with a mesenchymal phenotype, plasma cells, and B memory cells. Mg exhibited positive associations with B cell precursors and proliferating B cells. Mn was positively linked to dendritic cells. Zn and K both showed negative correlations with epithelial cells and red blood cells. Ca exhibited negative correlation with monocytes.

### Metals-Based Pathway Analysis

Our differential expression analysis uncovered a wide range of biological pathways associated with biomolecular accumulation, reflecting variations in cellular composition, immune responses, and tissue architecture (**Tables S4-7**). A demonstration of the spatial covariation between elements and specific genes can be found in **Figures 3,S1**. This analysis highlighted both shared and unique roles of different metals in key cellular processes, including but not limited to: *Immune Response, Inflammation, and Complement Activation.* We identified metals which co-occurred with genes involved in key immune signaling pathways such as Interferon Gamma Response (Cu, p=1.09e-12; Mg, p=6.46e-18; Mn, p=5.92e-18) and Allograft Rejection (Mn, p=1.38e-29; Mg, p=2.23e-22). Additionally, these metals were involved in Complement Activation and IL-2/STAT5 Signaling (Fe, p=0.000175), indicating their significant roles in modulating immune responses within the TME. Cu was also prominently linked to IL-6/JAK/STAT3 Signaling (p=0.00505), a pathway known to be involved in inflammatory responses and immune regulation. Moreover, Mg was associated with pathways like PD-1 Signaling (p=1.47e-06) and Phosphorylation of CD3 and TCR Zeta Chains (p=6.42e-05), further emphasizing the impact of these elements on immune cell activation and signaling.

**Figure 3:**
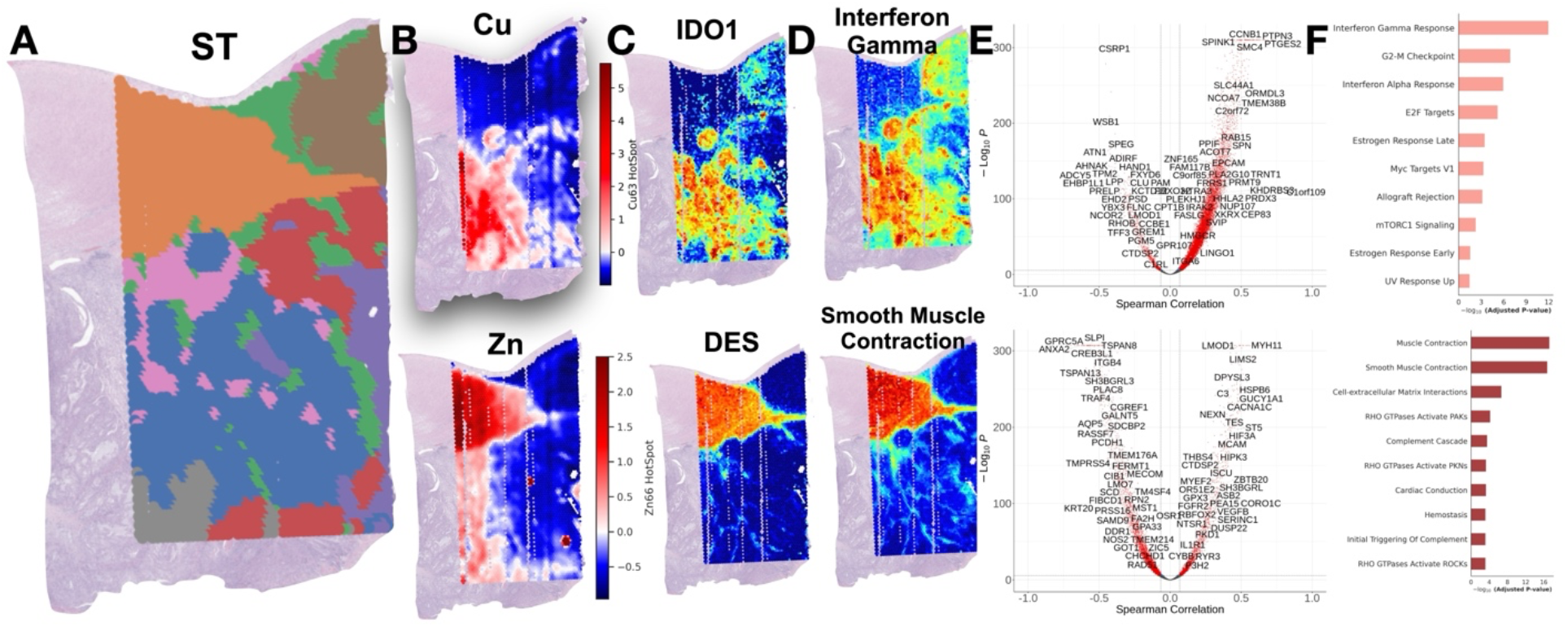
Pathway Analysis Findings: **A)** Visual representation of ST data through Leiden clustering, revealing distinct regions, **B)** Cu and Zn Gi* hotspot statistics, **C)** Visualization of spatial distribution of IDO1 and DES gene expression demonstrating association with respective elements, **D)** Visualization of Interferon Gamma Response and Smooth Muscle Contraction through aggregating gene expression across pathway compared to a background distribution, again found to be associated with respective elements, **E)** Volcano plot mapping spearman correlation between 18074 genes and Gi* statistics for respective metals on x-axis and -log10(p-values) on y-axis– p-value and correlation cutoffs were set at 0.05/18074; **F)** Pathways associated with respective elements (top includes most significant MsigDB Cancer Hallmarks for Cu abundance and bottom includes most significant Reactome pathways for Zn abundance)

*Epithelial-Mesenchymal Transition (EMT), Cell Motility, and Tissue Remodeling.* Fe, along with K and Zn, was prominently associated with pathways involved in EMT and cell motility, highlighting their roles in processes like EMT (Fe, p=4.38e-21) and Cell-extracellular Matrix Interactions (Fe, p=9.41e-11; Zn, p=2.06e-07). These elements were also linked to Apical Junctions and RHO GTPase activation, both of which are crucial for cell adhesion, migration, and tissue remodeling (Fe, p=2.75e-05; Zn, p=1.17e-06).

*Myogenesis and Muscle Contraction.* K and Zn were strongly associated with pathways related to myogenesis, muscle contraction, and ECM interactions, emphasizing their crucial roles in tumor progression. For example, myogenesis was significantly linked with K (p=5.48e-15).

Similarly, Smooth Muscle Contraction was associated with K (p=2.36e-15) and Zn (p=1.69e-17). Zn was also found to play a significant role in Muscle Contraction (p=5.79e-18).

*Cell Signaling, Apoptosis, and DNA Repair.* Pathways related to cell signaling, apoptosis, and DNA repair were notably enriched for elements such as Cu, Mg, Mn, and Fe. For instance, Cu showed significant enrichment in the G2-M Checkpoint (p=1.30e-07) pathways. These elements also demonstrated enrichment in apoptosis-related pathways (Mg, p=7.49e-03).

*Oxidative Stress, Cellular Repair, and Hormonal Regulation.* Fe was prominently associated with pathways related to oxidative stress, cellular repair mechanisms, and hormonal responses. Fe’s involvement in the Hypoxia pathway (p=0.000881) and Angiogenesis (p=0.0104) highlights its critical role in cellular adaptation to low oxygen levels, a hallmark of rapidly growing tumors. Additionally, Fe was strongly linked to KRAS Signaling Up (p=1.60e-16), reflecting its role in key oncogenic signaling pathways. These associations underscore the multifaceted importance of Fe in tumor growth, stress responses, and repair mechanisms within the TME. The depletion of pathways like and p53 Pathway (Ca, p=1.25e-08) may further indicate alterations in hormonal signaling and cellular stress responses within the TME.

*Metabolic Regulation and Signal Transduction.* Metabolic pathways, including glycolysis and cholesterol homeostasis, were frequently enriched in Zn and Cu, highlighting their roles in tumor metabolic adaptations. Zn was significantly involved in Glycolysis (p=7.09e-07), while Cu showed enrichment in mTORC1 Signaling (p=5.05e-03), emphasizing their contributions to the metabolic flexibility required for tumor survival within the TME.

### Further Examination of Cu Homeostasis and Fe-Related EMT Pathways

We further examined the relationship between specific genes and metal abundance within key pathways using PathVisio, overlaying correlations onto WikiPathway diagrams (**Figures S2-3**) ^49–54^. For genes correlating with Cu abundance, significant associations were observed within Cu homeostasis pathways. In particular, metal ion solute carrier (SLC) transporters MT1E (ρ=0.40, p<0.001) and MT1G (ρ=0.40, p<0.001) were strongly correlated with Cu levels.

Additionally, ATOX1 (ρ=0.15, p<0.001), a Cu chaperone delivering Cu+ to P-type ATPases such as ATP7A (ρ=0.31, p<0.001), exhibited notable correlations. Through its suppressive effects on SOD3, a key antioxidant gene, ATP7A may influence tumor progression ^55–58^. SOD3 protects against oxidative stress and maintains cellular redox balance, and its downregulation has been associated with increased oxidative stress and induction of EMT, processes linked to tumor metastasis ^59^. Correspondingly, SOD3 (ρ=-0.36, p<0.001) showed the strongest negative correlation with Cu abundance within this pathway, suggesting that elevated Cu levels may drive downregulation of this protective gene through upregulation of ATP7A, further promoting cancer aggressiveness.

For Fe abundance, spatial correlations were identified with specific genes linked to EMT-related pathways. Among these, FN1 (ρ=0.34, p<0.001), which encodes fibronectin, a protein involved in promoting cell motility through collagen matrix remodeling and serving as a marker for cancer-associated fibroblasts ^60,61^, was most notable. Additional key correlations were observed with SPARC (ρ=0.36, p<0.001), which phosphorylates focal adhesion kinase (FAK) to stimulate tumor cell invasion ^62^, and MMP9 (ρ=0.32, p<0.001), a matrix metalloproteinase associated with extracellular matrix (ECM) degradation, lymph node metastasis, and poorer survival outcomes ^63,64^. By examining these specific gene correlations within their respective pathways, our analysis highlights the intricate interplay between metal abundance and gene activity in the tumor microenvironment.

### Spatial Clustering of Metals, Genes, Cell-Types

The pathway associations identified in our analysis suggest significant roles for various metals in critical biological processes such as immune response, cell cycle regulation, and ECM interactions. However, these associations raise important questions about how these processes are spatially organized within the tumor microenvironment. Spatial colocalization of metals and specific cellular activities is likely more pronounced in certain regions of the tissue, where distinct tissue architectures may drive localized biological effects. To explore these spatial patterns and their implications for tumor behavior, we conducted a spatial factor analysis to summarize these associations within distinct tissue architectures (**Figure 4**).

**Figure 4:**
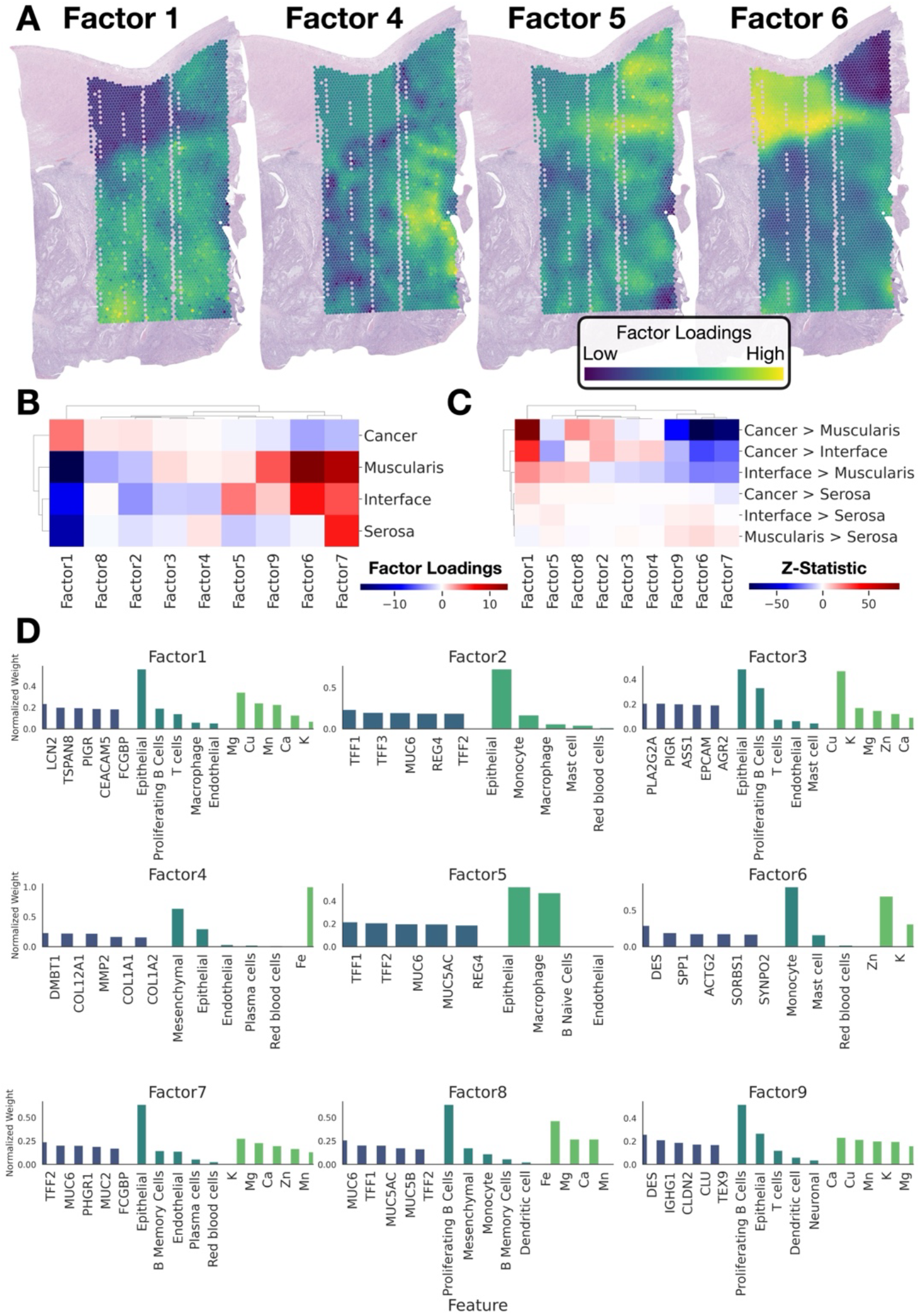
Spatial Multimodal Factor Analysis Results: **A)** Plotting of factor loadings from four of nine discovered spatial factors, **B)** Clustermap demonstrating factor loadings averaged across architecture, **C)** Relative differences in factor loadings between tissue architectures, hierarchically clustered, **D)** Composition of factors, top five features per data type (genes, cell types, metals); contributions of these features were normalized to sum to one within each data type

The spatial factor analysis using MEFISTO yielded nine factors corresponding to a variety of biological pathways (see **Figures S4-5, Tables S8-10**) ^65^. Factor 1 was primarily concentrated within the tumor and was associated with the accumulation of Mn and Cu, alongside pathways related to mucin O-glycosylation, glycolysis and estrogen response ^66–68^. Factor 4 was linked to a mesenchymal phenotype and associated with Fe, showing strong connections to gene signatures tied to EMT. Factor 5 was predominantly localized at the tumor interface, associated with both epithelial and macrophage cells, and was enriched in pathways related to EMT and immune responses, including Dectin-2 and Defective GALNT12 causes CRCS1. Finally, Factor 6 was connected to myogenesis, muscle contraction, and monocyte presence, with a concentration of Zn at the tumor interface and in the muscularis.

### How Much of the Spatial Elemental Distribution Can Be Explained by Transcriptomic, Cellular, and Pathway Activity Variation?

We used a multivariable predictive modeling approach with the MISTy package to evaluate how well transcriptomic, cellular, and pathway activity variations explain the spatial distribution of elements within the tumor microenvironment ^69^. This approach employed spatially weighted random forest models to predict metal concentrations based on pathway activity scores, cell type abundances, and the spatial gene expression of selected genes at the same or neighboring spots. Our analysis found that a substantial proportion of the variation in elemental distribution can be attributed to these spatial transcriptomic data types. For elements such as K, Cu, Mn, Mg, Ca, and Fe, the models achieved R² values exceeding 75%, indicating a high level of explainability (**Figure S6, Table S11**).

Among the predictors, spatial gene expression markers were the most significant influences upon elemental variation. In contrast, pathway activity scores and cell type abundances were less predictive, potentially due to the information loss during data aggregation. Interestingly, when incorporating other elemental concentrations as predictors, the majority of the variation in elemental distribution was accounted for by these other elements and cell types, rather than gene expression alone. A comprehensive list of gene, cell type, and pathway interactions with metals is provided in **Figure S6** and **Table S11**.

## Discussion

Targeting metal-dependent signaling and chaperoning within and around tumors has emerged as a promising strategy for inhibiting tumor growth and spread ^5,12,70^. However, this approach is challenging due to the limited understanding of how metals accumulate within tumors.

Developing a comprehensive map of the conditions and consequences of metal accumulation in tumors and their surrounding microenvironment is crucial for advancing next-generation cancer therapies. The differential bioaccumulation of endogenous metals, which may reflect variations in nutrient intake, storage, or disruptions in homeostasis, underscores the need for such a map. This metal map would provide valuable insights into the bidirectional molecular mechanisms governing metal accumulation, whether in excess or deficiency. By understanding how these metals contribute to tumor progression, we can better identify potential biomarkers and therapeutic targets, paving the way for more effective treatments.

In this study, we focused on spatially characterizing CRC tumorigenesis, as lifestyle factors such as dietary intake, which is one of many factors influencing metabolic activity and inflammation, can significantly impact tumor development, offering another promising therapeutic avenue in addition to chelation. The computational workflow that was developed and implemented is the first of its kind and has uniquely layered on spatial transcriptomics atop elemental imaging to facilitate a metals-based pathway analysis. Our workflow offers a preliminary understanding of dynamic interplay between metallic and molecular alterations within the TME.

By integrating histology imaging and pathology annotation, we demonstrated metal bioaccumulation across various tumor microenvironment compartments, with Cu localizing intratumorally and Fe accumulating at the tumor’s proliferative front and within the stromal architecture. Cu, essential for mitochondrial energy metabolism in cancer cells, was primarily localized within the tumor, corroborating prior studies showing that Cu chelation can kill colon cancer cells by preventing redox cycling and thereby reducing the generation of reactive oxygen species. Fe, on the other hand, was found in the tumor stroma ^71,72^. This finding aligns with prior research suggesting that Fe accumulation in this region may result from residual Fe following intralesional hemorrhage. Larger tumors, with their fragile neovascular blood vessels, are prone to bleeding, which could contribute to this stromal Fe deposition, as demonstrated through supporting literature ^73^. Indeed, deposits of Fe were found to be associated with hypoxia-related genes. These findings were further supported by spatial integration of single-cell data with ST to derive cell-type proportion estimates, which established an epithelial association for Cu and a mesenchymal/stromal phenotype for Fe. The presence of Fe in regions tied to stromal/collagen remodeling aligns with the epithelial-to-mesenchymal transition, a key process in tumor progression. Additionally, Cu was not only found within tumor areas but also co-localized with endothelial cells, plasma cells, and macrophages. Pathway analysis recapitulated Cu’s dual role: promoting CRC proliferation, as indicated by enrichment in Mitotic Cell Activity, and stimulating the antitumoral immune response through interferon signaling and immune recruitment via the surrounding vasculature and lymphatics, consistent with prior literature ^74^. Both Cu and Fe findings were reinforced by differential expression analyses and visualization of spatial metal-gene correlations overlaid on pathways governing Cu homeostasis, metabolic reprogramming, and Fe-related EMT processes. These pathways highlight where their shared contributions relate to aggressive tumor phenotypes. Future studies are needed to validate these associations and disentangle the roles of Cu and Fe within specific cell mixtures localized in distinct tissue architectures. Together, these findings provide a nuanced understanding of Cu and Fe’s roles in tumor progression, consistent with prior literature while demonstrating the potential for new insights spurred through spatial integration and pathway-level analyses.

While Cu and Fe are key metals in cancer biology, other metals, such as Mn and Mg, also showed significant tumor concentrations and warrant further investigation for their distinct roles. Neither Mg nor Mn was associated with an epithelial phenotype, suggesting their involvement with other tumor-infiltrating cells. Previous research has linked increased cellular Mg to DNA and protein synthesis, as well as tissue growth, which aligns with our observation of Mg’s localization within the tumor and its association with the G2M cell cycle pathway ^75^. Like Cu, Mg was also highly enriched for genes associated with inflammatory signaling via the Interferon/cytokine signaling pathway, among others. In contrast, Mn localized within the tumor likely due to its role in the antitumor immune response ^76^. Prior studies have suggested Mn’s role in promoting dendritic cells, which is consistent with our findings showing the strongest correlation of Mn with dendritic cell presence within the tumor.

Zn was found colocalized at the tumor interface and has been shown in prior studies to alter cytoskeletal integrity, motility, and invasiveness of colon cancer cells, suggesting a potential role in chemoprevention. This cytoskeletal activity is consistent with pathway activation related to myogenesis and muscle contraction, though not much is known about its precise mechanisms within tumors.

The presence of K and Ca at the tumor interface may reflect long-term accumulation in the colon’s muscularis propria, which consists of older, longer-lasting cells. This buildup is likely due to extended exposure to these elements. As a validation, we also found Ca ions within the same regions. Previous studies have shown that Ca affects intestinal smooth muscle contraction, partly by influencing its permeability—consistent with our findings of Ca within these architectures ^77^. Additionally, both Ca and K were related to Ca²⁺-activated K⁺ channels, supporting this effect and validating our approach.

These associations underscore the unique potential enabled by integrating these advanced technologies. It’s important to highlight that a significant proportion of elemental distribution within tumors has been confirmed to have biomolecular underpinnings that govern not only metal deposition, redistribution, and chaperoning but also the downstream consequences of metal presence, such as inflammation. Developing mechanistic associations between these findings and tumor biology will take time, but the progress is promising.

The advantage of pathway analysis through whole transcriptomic profiling lies in the broad scope and range of pathways that can be explored, offering real potential for biomarker and intervention discovery. However, it is crucial to emphasize that while some findings in this manuscript are confirmatory, they are largely exploratory and require validation and expansion to a larger, unbiased cohort. Some of the pathways identified in this analysis reflect normal colonic function rather than tumorigenesis. We have been careful to limit the set of considered elements to avoid overstating conclusions. Future work will delve deeper into cell-type associations using complementary imaging techniques and will also investigate alterations that exceed those attributable solely to tissue architectural changes. Key indicators of tumor progression and metastasis can only be fully understood when employing these technologies on a larger scale / expanded cohort. CRC progression and tumorigenesis are heavily influenced by various factors, including tumor sidedness, genetic and lifestyle factors, invasiveness, grade, sex, and other confounders and modifiers including deficiencies in mismatch repair was the case with this patient which can reflect a T-cell exhausted phenotype. Addressing these complexities will be essential in advancing our understanding and treatment of CRC.

Furthermore, tissue sections were separated by 5 microns, which assumes smooth tissue changes and may lead to potential imprecisions in co-registration, thereby impacting the findings. Profiling device noise also introduces additional challenges with regards to precision. Tissue was analyzed with paraffin intact which can lead to random signal attenuation due to build-up of paraffin within the LA-ICPTOF-MS capture tube if the capture tube is too small. We did not deparaffinize because it has the potential to shift elemental distribution and reduce abundance. There are also biological buffers employed at various parts of tissue preparation that could influence findings– we were careful to remove elements that could have been influenced by preparation or overly represent individual variation. In the future, increasing the size of the capture tube should significantly reduce impact of paraffin at the cost of resolution (i.e., from 1-micron to 5-10 micron resolution). In our integrative analyses, metal abundance was aggregated at a 50-micron resolution, which we felt was reasonable. Single-cell analysis was not possible at this time due to the 5-micron separation between EI and ST sections and the laser-destructive nature of this process– future work in this area may enhance the resolution of these findings and better appreciate tissue changes between adjacent sections ^78^. It should also be noted that due to the ionization of particles, LA-ICPTOF-MS was not developed to characterize valence states. However, further understanding of speciation components that catalyze biomolecular interactions may be accomplished by pairing this technique with others, such as X-ray absorption near-edge structure (XANES) ^79,80^. Analyzing smaller tissue regions at higher resolution may help deduce specific species, though performing these analyses at scale remains challenging.

This approach serves as a proof-of-concept workflow, demonstrating how the spatial integration of metals and gene analysis can soon be used to effectively capture the biological processes governing tumor metastasis, recurrence, and survival. This study paves the way for comprehensive exploration of spatial elemental data and gene expression in colorectal cancer and other tumor types, offering opportunities to advance therapeutic development, understand the biological and prognostic significance of elemental shifts, and investigate the impact of dietary intake on metal redistribution in early onset and progression of CRC.

## Methods

### Cohort Curation and Data Collection

In our study, we focused on a specific patient, selected randomly from a cohort of 45 specimens recently profiled using Visium Spatial Transcriptomics (ST). The selected patient, a 55-60 year-old female, had a left colon microsatellite stable (MSS) tumor (intact MLH1, MSH2, MSH6, and PMS2 expression assessed through immunohistochemistry) that metastasized to the liver. The tissue sections were processed using the Sakura Tissue-Tek Prisma Stainer for hematoxylin and eosin staining. For ST profiling, 5 μm tissue segments were sectioned and dissected from formalin fixed paraffin embedded tissue blocks, placed within a 11mm by 11mm capture region. A separate unstained 10 μm serial section was cut for elemental imaging. Finally, a subsequent 5 μm section was left intact without macrodissection and was stained with H&E. The slides were scanned at 40X magnification (approximately 0.25 micron per pixel) using the Aperio-GT450 scanner (Leica, Wetzlar, Germany). The resulting hematoxylin and eosin-stained images were stored in SVS format with eight-bit color channels.

### Elemental Imaging

For the elemental profiling of the colorectal cancer tissue section, we used Laser Ablation Inductively Coupled Plasma Time-of-Flight Mass Spectrometry (LA-ICPTOF-MS) ^81^. This method represents a significant advancement in spatially resolved elemental imaging, offering both enhanced resolution and analytical speed. In this process, a pulsed laser is used to ablate minute portions of the tissue sample. The ablated material, now in particulate form, is then carried via a helium gas stream into the mass spectrometer. The key feature of LA-ICPTOF-MS is its use of Time-of-Flight (TOF) technology, enabling rapid and comprehensive elemental detection across the entire periodic table. Operating at high frequencies (500-1000 Hz), the LA- ICPTOF-MS system at the Biomedical National Elemental Imaging Resource (BNEIR) allows for detailed mapping of elemental distribution with ultra-high resolution, down to 1 µm. This capability is crucial for accurately capturing the complex elemental landscape within the tissue, providing insights into the spatial relationships and concentrations of various elements. By utilizing LA-ICPTOF-MS, we were able to conduct an untargeted yet thorough profiling of the tissue section, yielding detailed data on its elemental composition.

### Spatial Transcriptomic Profiling and Spot-Level Cell-Type Deconvolution

We utilized the 10X Genomics Visium CytAssist spatial transcriptomics (ST) assay for in-depth profiling of a tissue section ^82^. The preparation of FFPE tissue sections involved several steps: firstly, placing the sections on standard histology slides, coverslipping in glycerol + xylene medium, followed by deparaffinization, rehydration, and H&E staining using a Sakura Tissue-Tek Prisma Stainer (Sakura Finetek USA, Inc. 1750 West 214th Street, Torrance, CA 90501). Subsequently, whole slide images were captured at 40x resolution on Aperio GT450 scanners (see *Cohort Curation and Data Collection*). The slides were then decoverslipped in xylene over 1-3 days to detach the coverslips. The subsequent steps, including destaining, probe hybridization, ligation, eosin staining, transfer to Visium slides via CytAssist, and library preparation, adhered to the manufacturer’s protocol (CG000485). After initial preparation, the tissue underwent eosin staining and imaging, aligning with the original high-resolution pathology slides. Tissue permeabilization followed to release mRNA molecules, which bound to target probes on the slide via their poly(A) tails. The binding process was succeeded by ligation, extension, and amplification of these probes. Sequencing was performed using an Illumina sequencer (NovaSeq 6000), enabling high-resolution gene expression mapping. The Spaceranger software was employed for precise alignment of the CytAssist sections with the corresponding 40X H&E stains, ensuring accurate co-registration and quality control.

Pathologists annotated the tumor’s interior, periphery/interface, and surrounding architectures using the QuPath tool, permitting delineation of transcriptomic profiles in various tissue regions. Label propagation was used to refine unassigned annotations. To further characterize the tissue architecture, Visium data underwent dimensionality reduction via UMAP projection, which served as a precursor to graph-based Leiden clustering ^83,84^. Subsequent refinement of cluster assignments was conducted through label propagation based on the spatial coordinates, specifically targeting areas with high entropy to enhance spatial consistency of the cluster labels. Finally, the delineated clusters were superimposed onto the whole slide images, with labels assigned in accordance with the tissue architecture as judged by a pathologist.

To understand the cellular composition within the CRC tissue, we combined spatial transcriptomics with single-cell RNA-Seq data collected from a serial section. We utilized the Chromium Flex assay for single-cell profiling of disaggregated FFPE tissue sections from specific capture areas, employing the same transcriptomic probe set as the Visium assay. This approach revealed diverse cell types within the tissue, following the manufacturer’s Demonstrated Protocol (CG000606). The generated data, processed using CellRanger v7.1.0, provided quality control metrics and cells-by-genes expression matrices for downstream analysis. For label transfer, cell types from a public single-cell RNA sequencing (scRNA-seq) dataset ^85^, specifically categorized cells from the Colon, were used. Cell types were grouped into broader categories. Dendritic Cells encompassed cDC1, cDC2, Lymphoid DC, and pDC cells. Epithelial Cells, which in this context are representative of tumor cells, also leveraged signatures defining Goblet cells, Colonocytes, and Enterocytes, and so forth. Endothelial Cells were grouped to cover a range of arterial, venous, and lymphatic subtypes. Macrophages were categorized based on LYVE1+ and MMP9+ subtypes, while Mast Cells included both Mast cells and CLC+ Mast cells. Mesenchymal Cells, representing the tumor and normal fibrous stroma, included stromal cells, myofibroblasts, and pericytes. T Cells aggregated across CD4, CD8, and NK lineages. Plasma Cells included IgA and IgG subtypes. Additionally, Neuronal Cells, Monocytes, Neutrophils, Megakaryocytes, B Cell Precursors (including Immature B, Pro-B, Pre-B, and CLP cells), B Memory Cells, Proliferating B Cells and B Naive Cells, and Red Blood Cells were each treated as distinct categories. These cell type labels were transferred to our in-house CRC scRNA-seq dataset collected from a serial tissue section using SCVI (Single-Cell Variational Inference) framework ^86,87^, which leverages a denoising variational autoencoder (VAE) trained to infer cell types. We employed the Cell2Location package for spot-level deconvolution, using the scRNA-seq data as a reference to estimate cell-type proportions/abundances in each spot ^88^. This regression-based approach enabled us to spatially map the distribution of cell types within the CRC tissue section, yielding aggregate spot wise cellular abundances.

*Quality Control and Co-Registration via TRACE:* Recently, our team developed TRACE, a software tool under the Biomedical National Elemental Imaging Resource, specifically designed to co-register highly multiplexed elemental assays with tissue slides ^41,46,47^. TRACE enables comprehensive spatial multimodal tissue analysis by integrating spatial elemental and transcriptomic data. In this colorectal cancer study, TRACE was instrumental in co-registering multi-channel elemental images with whole slide images (WSI). Initially, TRACE’s preprocessing workflow aggregated elemental abundance across channels and user defined thresholding through interactive segmentation of background regions to accurately detect tissue in the elemental maps. Refinement of tissue detection involved Gaussian filtering for reduction of outliers, which can impact tissue detection, and morphological operations (binary opening/closing) to further refined these images, focusing on removing noise and defining contiguous regions for analysis (**Figure S7A**).

A key challenge in preprocessing was addressing directional stripe artifacts in elemental imaging. To tackle this, we employed anisotropic diffusion filtering to reveal sharp edge patterns in the tissue followed by the probabilistic Hough transform which is a line fitting and detection for isolating detected strips ^89,90^. Line detection was followed by morphological transformations to remove the artifacts (**Figure S7B-F**). This meticulous process ensured elemental maps were free of distortions that could impact the analysis. We also encountered metal washout at tissue edges, particularly for elements like iron, and implemented a targeted erosion technique to address this issue. This strategy selectively eroded the edges of the tissue mask, effectively minimizing edge-related distortions and ensuring a more accurate representation of elemental distribution within the core tissue areas (**Figure S7G**).

For co-registration, a landmark-based approach with Homography matrix transformation aligned the elemental maps with the ST-associated WSI. Selecting 30 manual fiducials that marked structural similarities between the elemental maps and WSI enabled us to precisely overlay the elemental map onto the WSI. Recall that ST, single cell data and pathologist annotations had been mapped to these same H&E WSI, providing a common reference frame for integration with the elemental maps. TRACE exports a SpatialData file containing elemental abundance and tissue region annotations supplied and integrated using QuPath ^91,92^. Using the Nearest Neighbors algorithm (capturing adjacent pixels within radius of Visium spot), we assigned pixel coordinates from the elemental maps to the nearest Visium spots. Elemental pixel values were aggregated within spot, combining elemental and transcriptomic data onto a unified frame using SpatialData (interoperable with Anndata and Muon) data formats for in-depth analysis ^93^.

### Hotspot Analysis

We represented the spatial distribution of elemental metal concentrations within tissue sections by identifying areas of concentrated metal abundance. This was achieved by calculating hotspots using the Getis-Ord Gi* statistic ^48^, implemented through the pySAL package ^94^. The Getis-Ord Gi* statistic, a spatial statistic, evaluates the metal concentration in each pixel or spot in relation to its neighbors (taken to be spots within approximately 80 micron), producing spot-level z-scores and p-values. These scores helped us discern statistically significant areas where metal concentrations were either unusually high (hot spots) or low (cold spots) compared to the expected local average. Our approach involved permutation testing and normality assumptions to ensure the robustness of identified hotspots. Throughout the remainder of the manuscript, we used Gi* z-scores to represent elemental hotspotting, serving as the primary variable to reflect elemental abundance.

### Association of Hotspots with Tissue Histology and Cell-Type Abundances

Linear regression models were used to associate the hotspot Gi* z-scores representing elemental abundance (dependent variable) with specific tissue architectures (four regions– cancer, serosa, interface, muscularis) represented as categorical fixed effects. Post-hoc pairwise comparisons between tissue architectures (e.g., tumor vs. interface) were conducted using estimated marginal means (R v4.1, emmeans package), which calculated mean Gi* statistics (hotspot concentrations) for each tissue type/architecture ^95^. The mean statistics and their relative differences estimated through the linear modeling were hierarchically clustered to reveal similarities between elements in their distribution. Similarly, spatially-integrated cell-type proportions were associated with varying metal concentrations represented using Gi* statistics using spearman correlations. The spearman correlation matrix between metals and cell-types were hierarchically clustered to reveal metals with similar cell-type associations. Differences were visualized with boxplots, dotplots and clustered heatmaps with dendrograms.

### Differential Expression and Pathway Analysis

Spearman correlations were employed to compare spatial gene expression with elemental abundance, linking Gi* z-scores representing transformed elemental abundance as our elemental features with spatial transcriptomics across the entire transcriptome for each metal. Results were visualized using volcano plots ^96^. After Bonferroni adjustment to account for multiple comparisons (alpha significance level of 0.05/18074 for 18074 genes tested), we selected the 150 top differentially expressed genes for each element, ranked by their adjusted p-values (all 150 genes surpassed the Bonferroni adjustment for all metals). Separate sets of 150 genes were selected based on whether correlations were positive, negative or based on the magnitude of the correlation. For each metal, pathway analyses using the Enrichr package were conducted across various pathway databases (including MsigDB Cancer Hallmarks 2020, Reactome 2022) for genes associated with positive and negative elemental concentration changes, and separately for genes regardless of whether their test statistics were positive or negative ^97^. Using PathVisio, metal gene correlations for Cu and Fe were also visualized atop genes involved in Copper Homeostasis and Epithelial-to-Mesenchymal related pathways, downloaded from WikiPathways ^49–54^.

### Integrating Spatial Multimodal Analysis to Profile Elemental, Genetic, and Cellular Co- Localization and Their Interactions

MEFISTO is a computational approach for analyzing multi-modal spatial biological data, unraveling latent factors that capture spatial variations linked to genes, metals, and cell types ^65^. It leverages tensor factorization alongside spatial and hierarchical Gaussian Processes to handle spatial autocorrelation, uncovering profiles that include gene expressions, elemental concentrations, and cell-type distributions. To refine the model, we limited factors to at most 20 factors, employed spike-and-slab priors for sparsity, and set Gaussian likelihoods for each data type with pseudo-log transformation for elemental abundances and cell type abundances, and Gi* z-scores for EI. For efficient training, we used a fast convergence mode leveraging sparse Gaussian processes, discarded low-impact factors, and harnessed GPU acceleration. Post-training, we examined the factors associated with specific tissue architectures using linear modeling on the spot-level factor loadings, similar to the above, and conducted a pathway analysis (Enrichr; MsigDB Cancer Hallmarks 2020, Reactome 2022) based on factor associated genes with an FDR adjusted p-value less than 0.05.

Unlike MEFISTO, which focuses on unraveling latent factors in multi-modal data, MISTy analyzes the spatial interactions among different features like genes, elements, and cell types. MISTy employs a multivariable model to determine how features including pathway activities, genes, and cell types relate to elemental abundance, using a spatially weighted approach ^69^. Specifically, it assesses the predictiveness of these factors on elemental abundance, employing a squared exponential decay kernel to account for spatial proximity. Pathway activity was estimated using the Progeny database through the Decoupler package ^98,99^. Spatially-informed random forest models were fit to predict elemental abundance from the spatial transcriptomic data types. The ability of the ST datatypes to explain elemental variation throughout the slide was estimated through the calculation of an element-specific R² gain score. MISTy broke this score down by each of the ST datatypes (scRNASeq/cell proportions, ST, pathway activity), representing their relative contributions. Contributions from other elements were included in the model (other than the one being estimated)– a new R² gain score was estimated which demonstrated the relative contributions of ST data versus the mixture/associations with other elements. Predictive elements, genes, pathways and cell-types were provided using feature importance scores via the impurity-based Gini index.

## Post-Hoc Exclusion of Elements and Isotopes

All detectable elements underwent spatial multimodal analysis. However, certain elements and select isotopes were removed post-hoc to ensure the reliability and biological relevance of the findings. Specifically, elements such as Na, Cd, Cr, V, As, Mo, Gd, Ru, Pt, Al, Ag, Se, Pb, Ba, Ni, Sb, Co, and Tl were excluded, despite their detection across the slide. These elements were removed to account for potential influences from tissue preparation processes, biological buffers, or individual variation, which could disproportionately impact their representation.

Additionally, some of these elements, while detected, were not previously hypothesized to exist at significant levels in the colon. Although their presence here is informative and warrants future exploration, we chose to exclude them to avoid overstating conclusions or misinterpreting findings in the current study. This selective approach ensures the remaining set of elements aligns with the study’s focus on tumor-related elemental interactions and provides a foundation for more targeted analyses in future work.

## Data Availability

Access to manuscript data is limited due to patient privacy concerns. All data produced in the present study are available upon reasonable request. Requests should be directed to senior author Dr. Joshua Levy.

---

**Figure S1:**
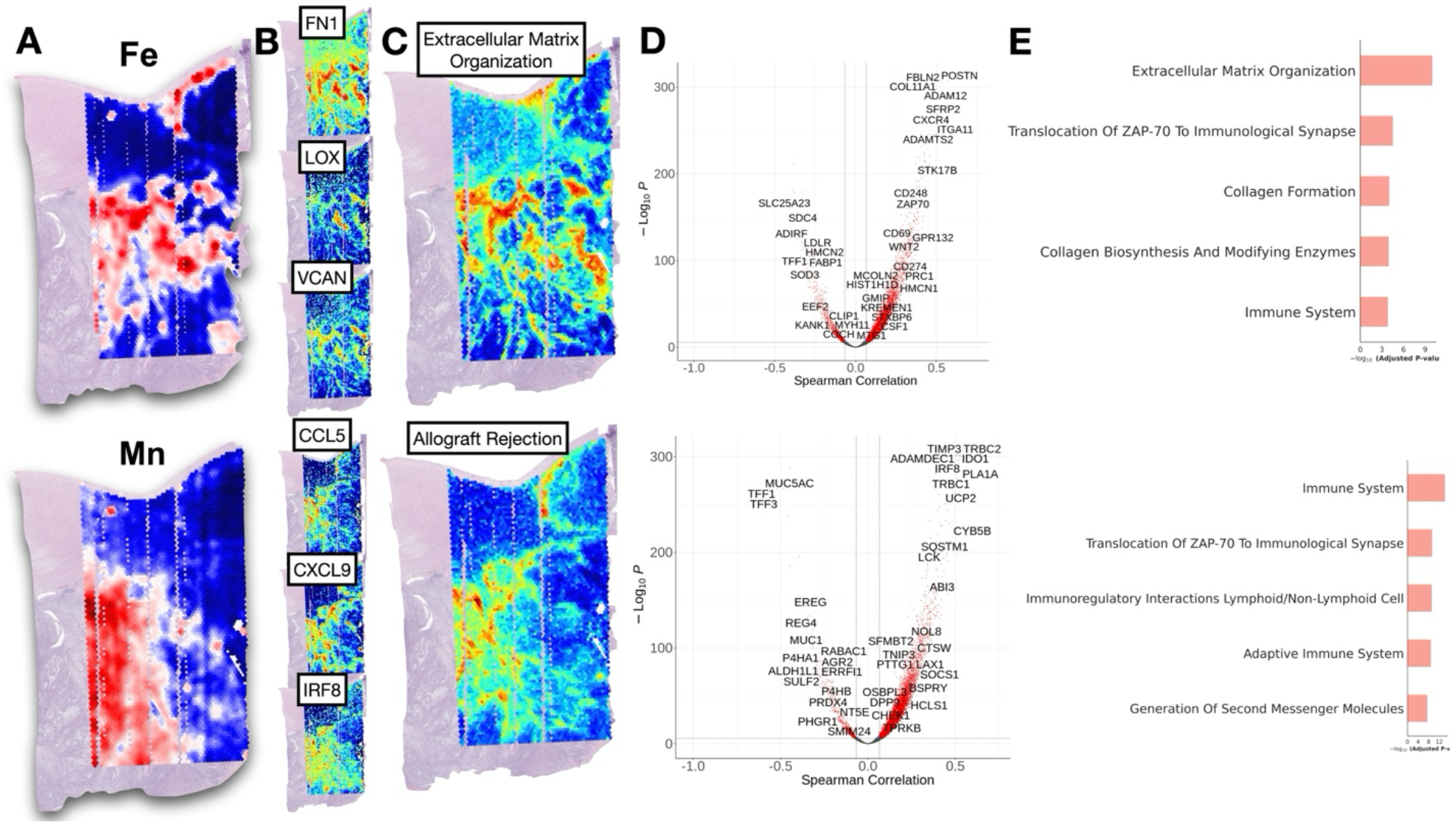
Additional Metals-Based Pathway Findings Based on Spatial Transcriptomics Correlation with Fe and Mn Distribution: **A)** Fe and Mn Gi* hotspot statistics, **B)** Visualization of spatial distribution of Fe-related (FN1, LOX, VCAN) and Mn-related (CCL5, CXCL9, IRF8) gene expression demonstrating association with respective elements, **C)** Visualization of Extracellular Matrix Organization and Allograft Rejection (reflects anti-cancer immune response) through aggregating gene expression across pathway compared to a background distribution, again found to be associated with respective elements, **D)** Volcano plot mapping spearman correlation between 18074 genes and Gi* statistics for respective metals on x-axis and -log10(p- values) on y-axis– p-value and correlation cutoffs were set at 0.05/18074; **E)** Pathways associated with respective elements (Reactome pathways)

**Figure S2:**
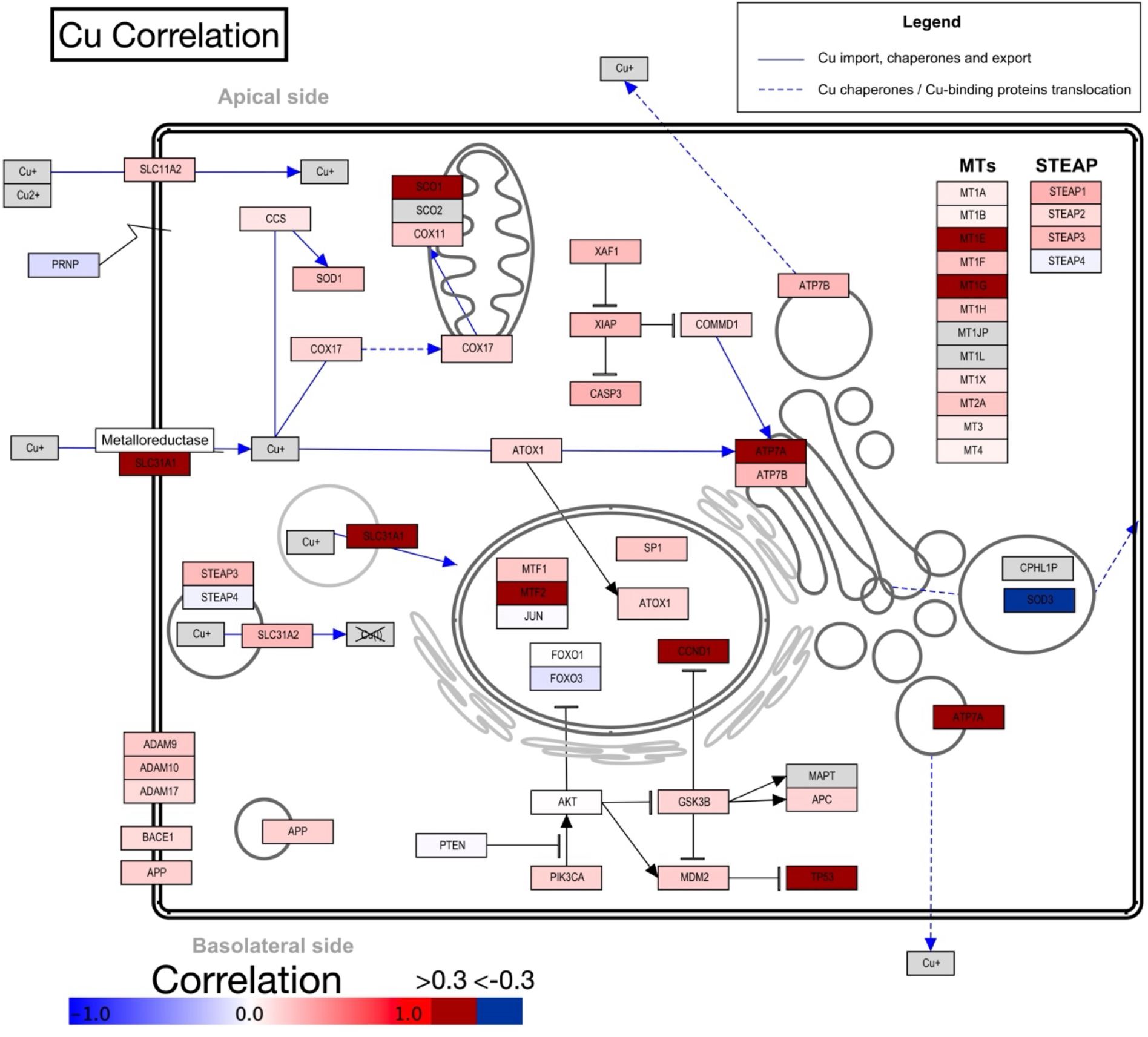
Visualization of Cu Differential Expression Results Overlaid on Cu Homeostasis Pathway Diagram: Color of each gene reflects positive (red) and negative (blue) correlations with Cu distribution; correlations with magnitude exceeding 0.3 are denoted using dark red/blue

**Figure S3:**
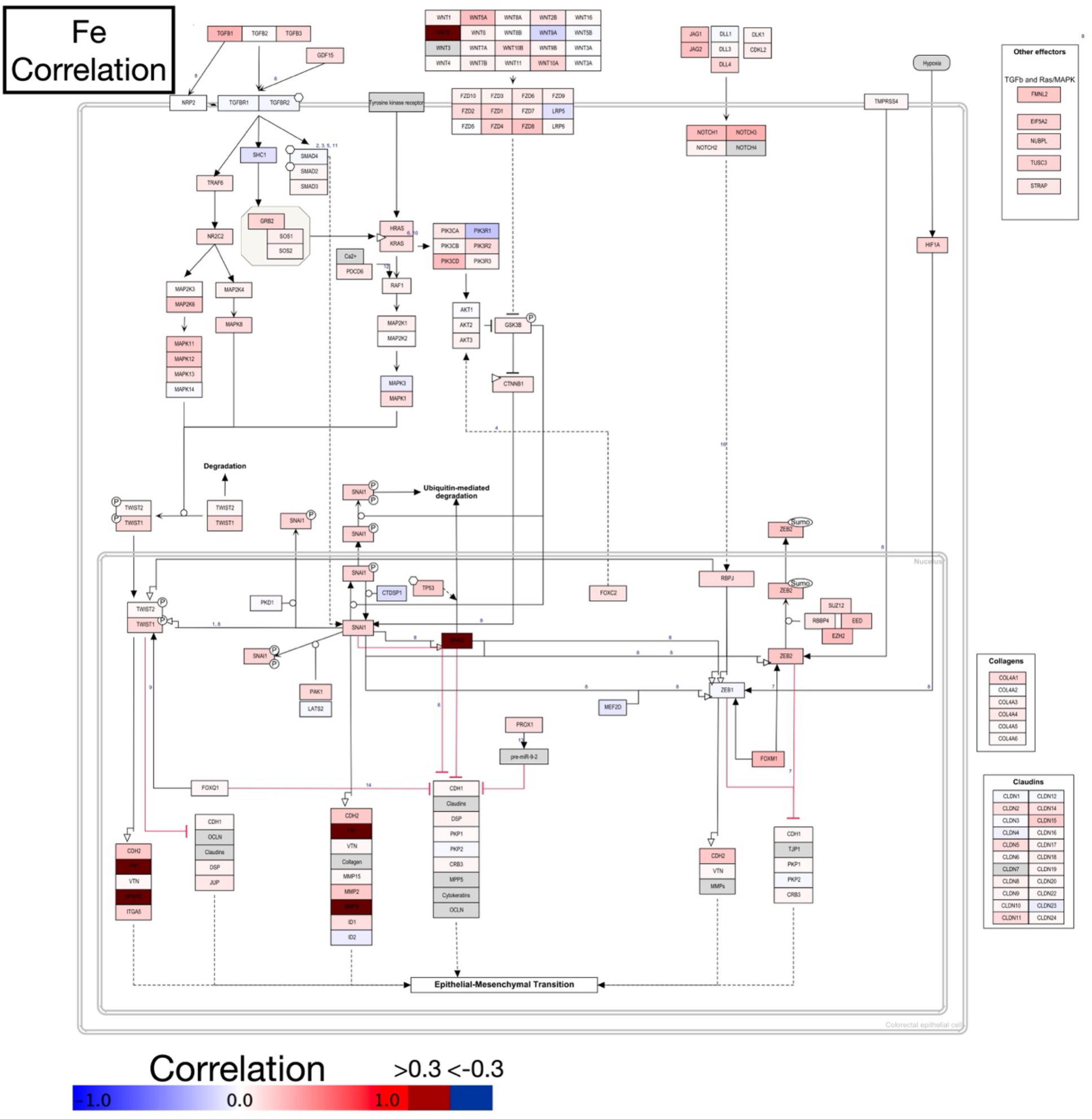
Visualization of Fe Differential Expression Results Overlaid on Epithelial-to-Mesenchymal Pathway Diagram: Color of each gene reflects positive (red) and negative (blue) correlations with Cu distribution; correlations with magnitude exceeding 0.3 are denoted using dark red/blue

**Figure S4:**
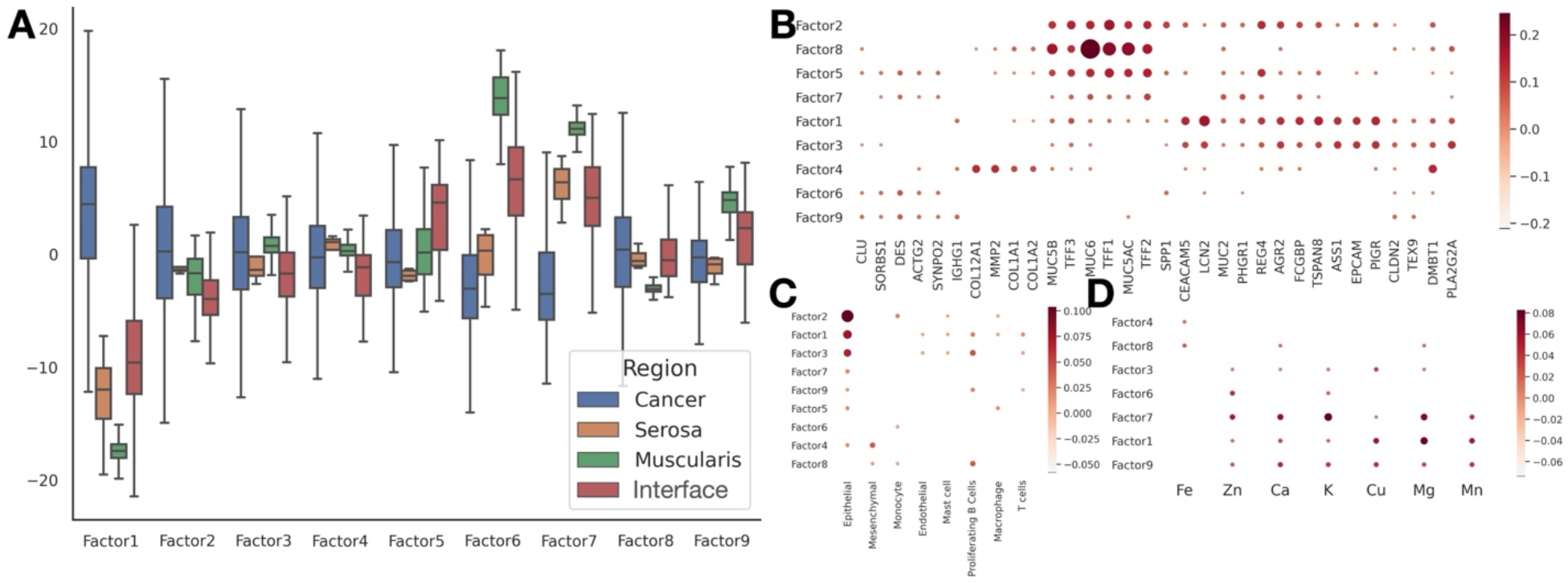
Further Factor Analysis Associations, broken down by: **A)** Tissue architecture (average loading), **B)** Gene expression (factor coefficients), **C)** cell type (factor coefficients), **D)** element (factor coefficients)

**Figure S5:**
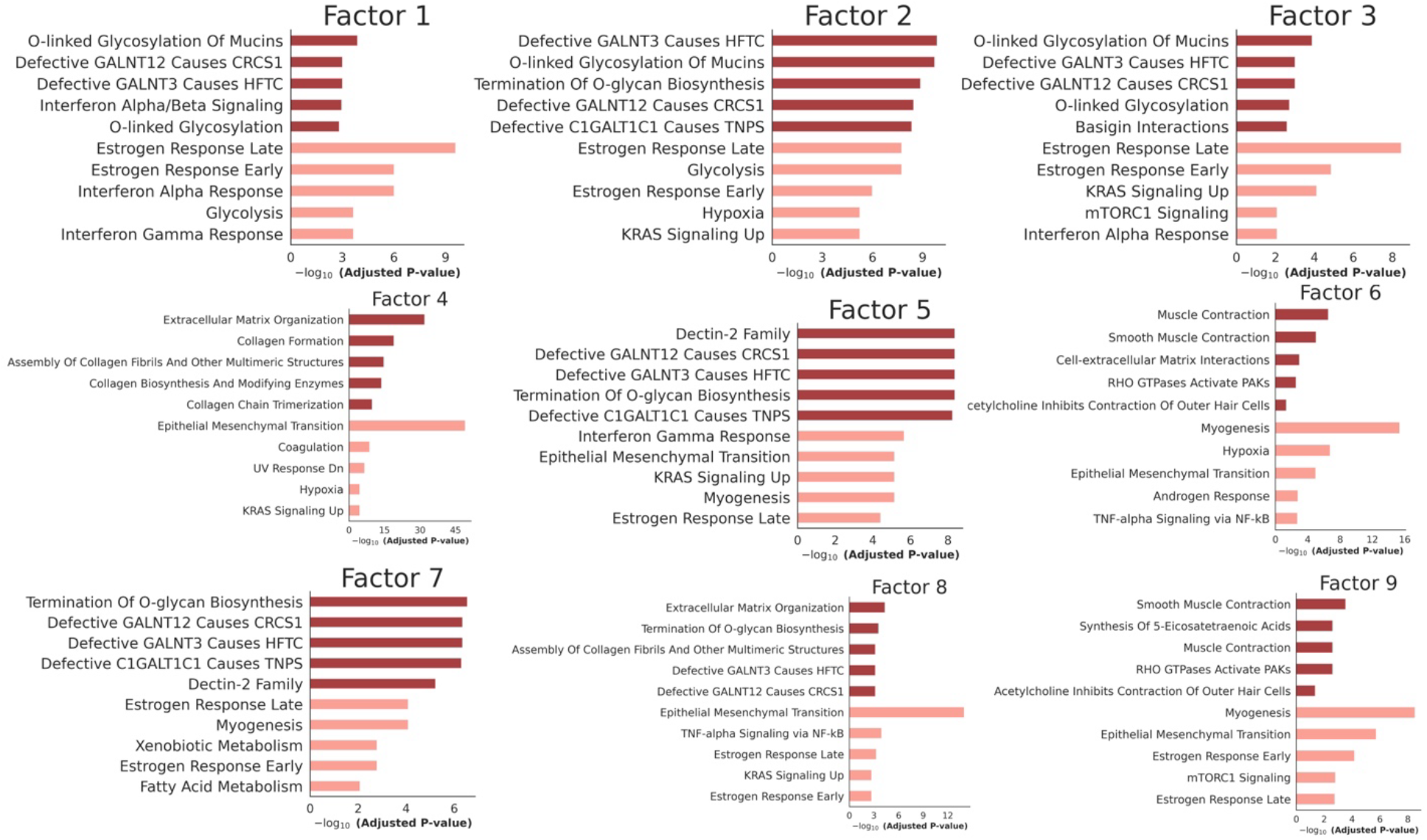
Association of Spatial Factors with Biological Pathways from Reactome (Dark Red) and MsigDB Cancer Hallmarks (Salmon)

**Figure S6:**
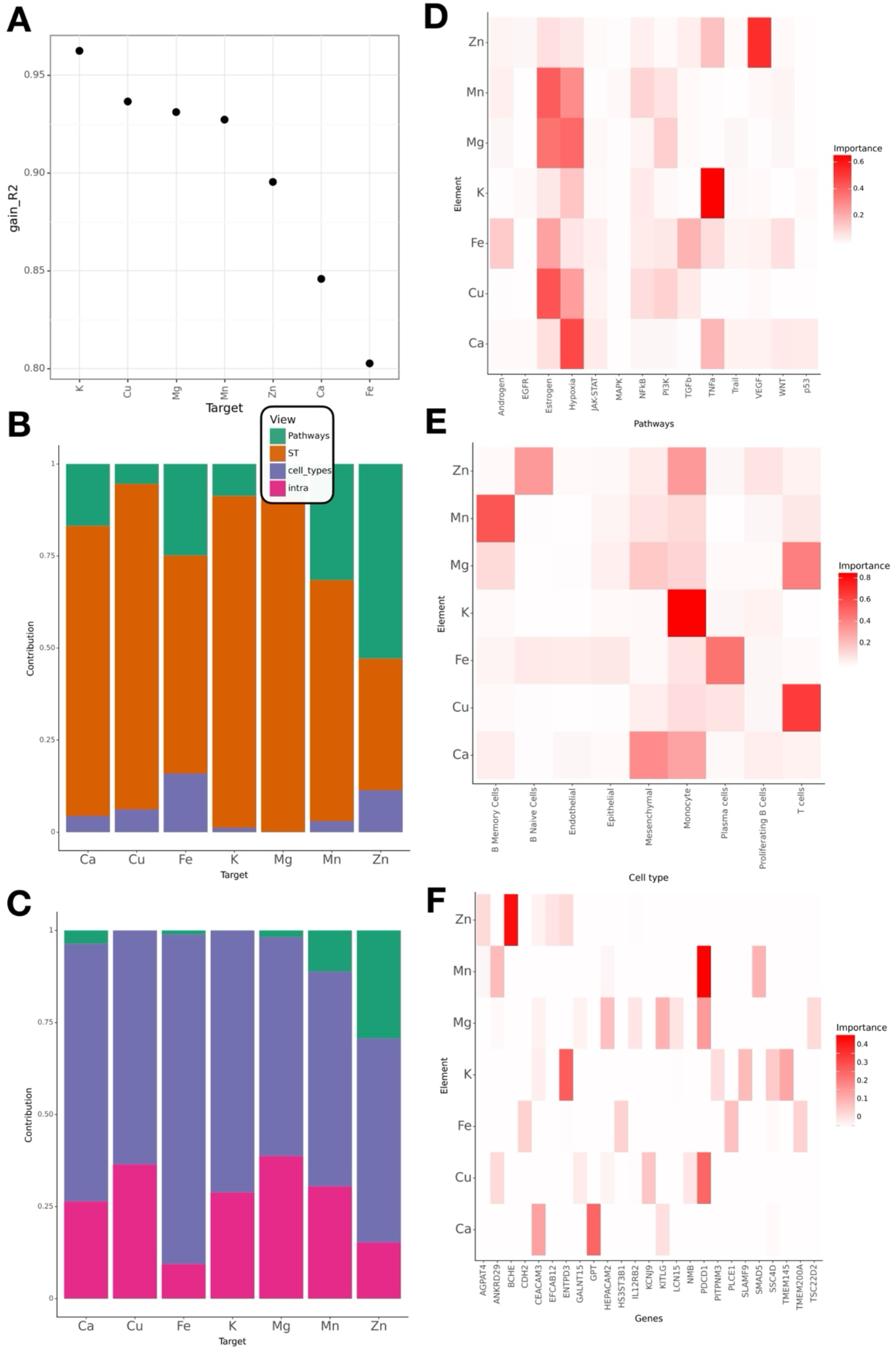
Elemental Distribution and Variance Explained via MISTy: **A)** Ability to predict various elements by ST data types alone via R², **B)** Breakdown of R² by data type, **C)** Breakdown of R² by data type when including other elements, **D-F)** Spatially informative features, specific to each element, broken down by **D)** Pathway, **E)** Cell type, **F)** Gene

**Figure S7:**
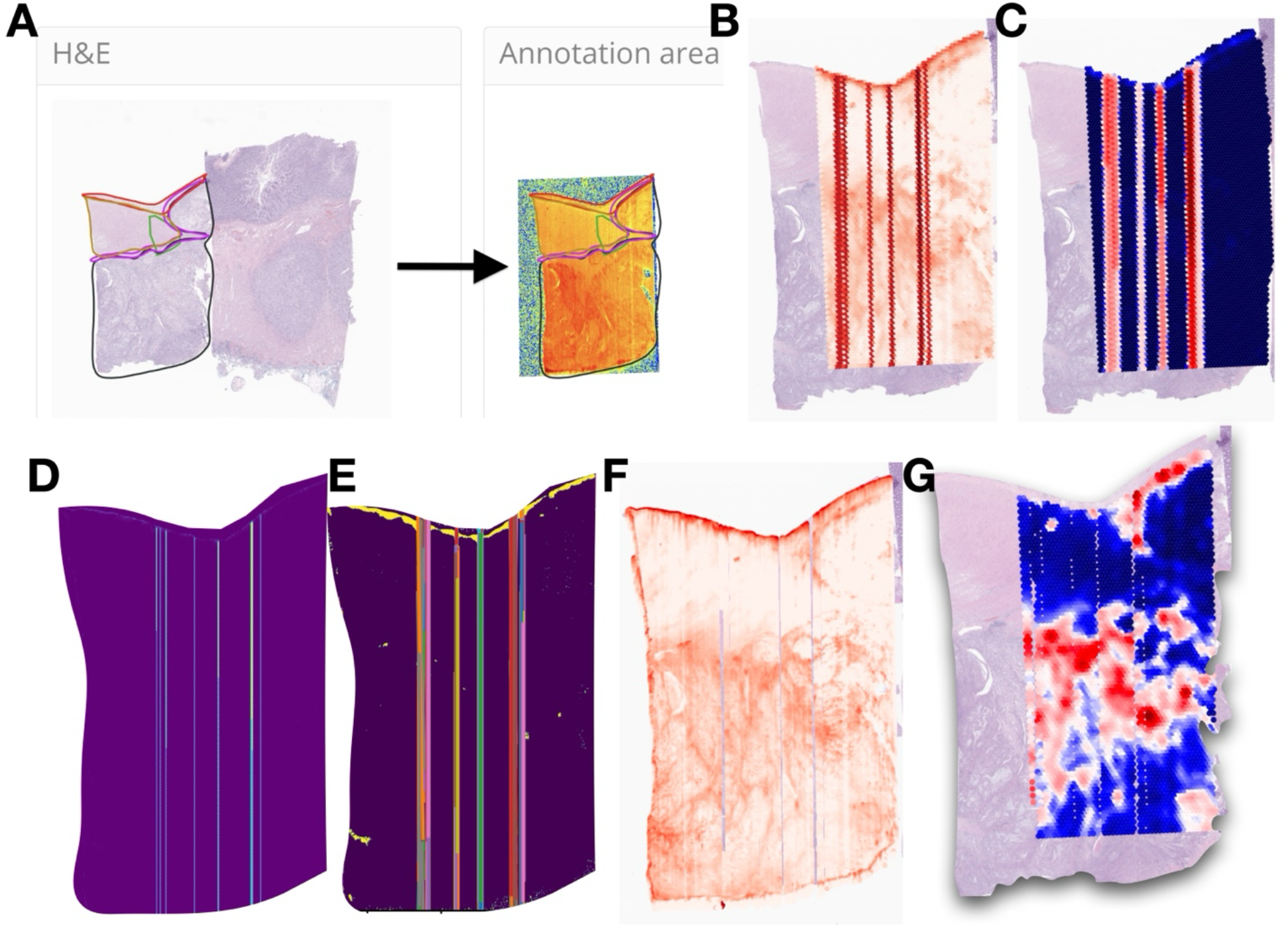
Preprocessing Workflow: **A)** Elemental maps are co-registered to ST-associated H&E WSI via TRACE, **B)** Vertical strip artifacts in Fe are **C)** confounding hotspot analysis and they are removed through **D)** anisotropic diffusion filtering, followed by **E)** hough transform for line detection. **F)** Elemental image after removing strip artifact. **G)** Removal of edge bleeding/washout artifact through binary erosion, aggregation of elemental abundance to ST spot level, and derivation of Gi* hotspot statistics

## Supplementary Tables

Table S1: Association of elemental abundance with architecture (one versus rest), includes average G* spot-level statistics by architecture and marginal mean calculated through regression modeling along with statistical significance (see TableS1.xlsx)

Table S2: Association of elemental abundance with pairwise relative differences between architectures, includes marginal mean comparisons calculated through regression modeling along with statistical significance (see TableS2.xlsx)

Table S3: Association of elemental abundance with cell-type, includes spearman correlations and p-values (see TableS3.xlsx)

Table S4: Differential expression analysis results, includes spearman correlation, p-values and Bonferroni-adjusted p-values by Element (see TableS4.xlsx)

Table S5: Pathway analysis findings (Reactome, Hallmarks) for top 150 genes tied to higher elemental abundance, ranked by z-statistic (see TableS5.xlsx)

Table S6: Pathway analysis findings (Reactome, Hallmarks) for top 150 genes tied to lower elemental abundance, ranked by z-statistic (see TableS6.xlsx)

Table S7: Pathway analysis findings (Reactome, Hallmarks) for top 150 genes tied to higher and lower elemental abundance, ranked by magnitude of z-statistic (see TableS7.xlsx)

Table S8: Composition of spatial factors, MEFISTO factor analysis weights by top 10 genes, cell-types and elements (see TableS8.xlsx)

Table S9: Association of spatial factors with architecture (one versus rest), includes averaged factor loadings by architecture and marginal mean calculated through regression modeling along with statistical significance (see TableS9.xlsx)

Table S10: Association of spatial factors with pairwise relative differences between architectures, includes marginal mean comparisons calculated through regression modeling along with statistical significance (see TableS10.xlsx)

Table S11: MISTy performance statistics and feature importances, includes metal abundance prediction performance broken down by data type (cell type information, spatial transcriptomics, pathways, metals/intra-view) with and without inclusion of other metals in model and Gini-index of features by data type (see TableS11.xlsx)

